# The purine pathway in liver tissue biopsies from donors for transplantation is associated to immediate graft function and survival

**DOI:** 10.1101/19005629

**Authors:** Jin Xu, Mohammad Hassan-Ally, Ana María Casas-Ferreira, Tommi Suvitaival, Yun Ma, Hector Vilca-Melendez, Mohamed Rela, Nigel Heaton, Wayel Jassem, Cristina Legido-Quigley

## Abstract

**Background & Aims:** The current shortage of livers for transplantation has increased the use of organs sourced from donation after circulatory death (DCD). These organs are prone to higher incidence of graft failure, but the underlying mechanisms are largely unknown. Here we aimed to find biomarkers of liver function before transplantation to better inform clinical evaluation.

**Methods:** Matched pre- and post-transplant liver biopsies from DCD (n=24) and donation after brain death (DBD, n=70) were collected. Liver biopsies were analysed using mass spectroscopy molecular phenotyping. First, a discrimination analysis DCD vs DBD was used to parse metabolites associated to DCD. Then a data-driven approach was used to predict Immediate Graft Function (IGF). The metabolites were tested in models to predict survival.

**Results:** Five metabolites in the purine pathway were selected and investigated. The ratios of: adenine monophosphate (AMP), adenine, adenosine and hypoxanthine to urate, differed between DBD and DCD biopsies at pre-transplantation stage (q<0.05). The ratios of AMP and adenine to urate also differed in biopsies from recipients undergoing IGF (q<0.05). Using random forest a panel composed by alanine aminotransferase (ALT) and AMP, adenine, hypoxanthine ratio to urate predicted IGF with AUC 0.84 (95% CI [0.71, 0.97]). In comparison AUC 0.71 (95%CI [0.52, 0.90]) was achieved by clinical measures. Survival analysis revealed that the metabolite classifier could stratify 6-year survival outcomes (p = 0.0073) while clinical data and donor class could not.

**Conclusions:** At liver pre-transplantation stage, a panel composed of purine metabolites and ALT in tissue could improve prediction of IGF and survival.

**Lay summary:** New liver function biomarkers could help clinicians assess livers before transplantation. Purines are small molecules that are found in healthy livers, and in this work we found that their levels changed critically in livers from cardiac death donors. Measuring them before transplantation improved the prediction of the liver’s immediate graft function.

**Graphic abstract:** 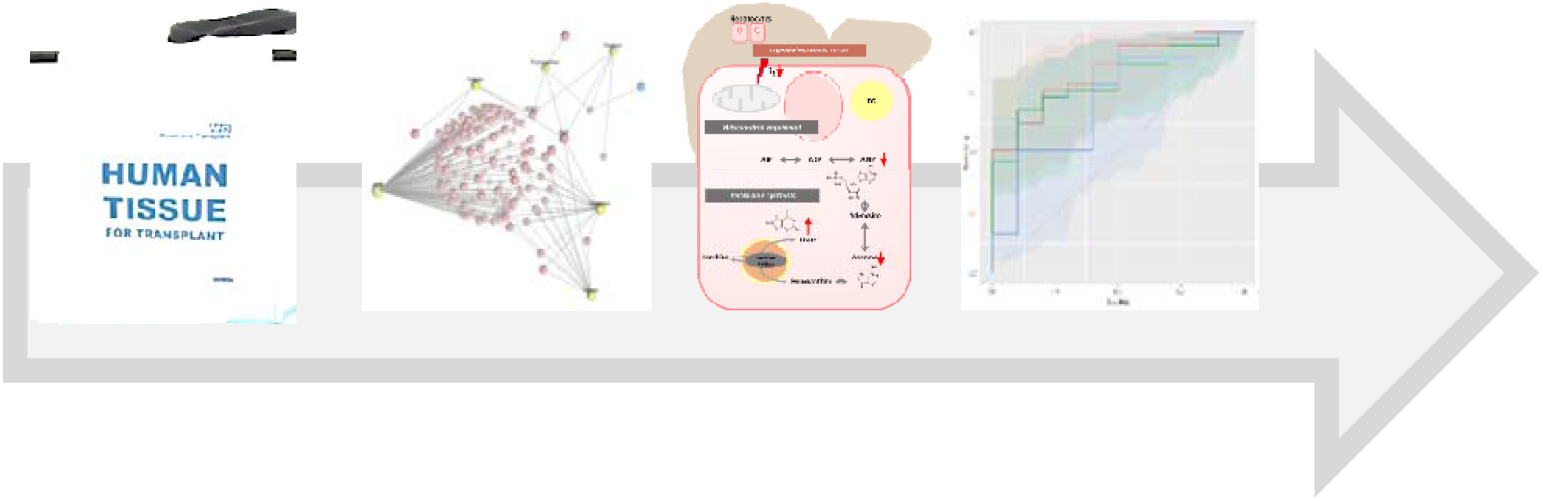

**Highlights:**

- The ratios of purine metabolites to urate differ between DCD and DBD in liver tissue at pre-transplantation.
- The ratios of purine metabolites to urate and ALT pre-transplantation can improve prediction of IGF after transplantation.
- Purine metabolites ratios to urate stratified 6-year survival outcome better than clinical data and donor class.

## Introduction

There is an ever increasing need for organ transplantation but the number of organs available is not increasing [1]. This is reflected by the number of people registered on the Organ Donor Register (ODR) in the UK which only saw a 5% rise from 2012 to 2017 [2]. While in the same period, the number of patients requiring a liver transplant increased from 452 to 519 [2]. This stark surge in the demand for liver transplants is attributable to the global incidence of liver drug-induced hepatotoxicity, fatty liver disease, cirrhosis and hepatitis infections [3].

To safeguard patients, pre-transplant donor screening is used to determine the success probability of liver transplants. The optimal/sub-optimal criteria for liver donors include age (<50 years/>50 years), weight (<100kg/>100kg), intensive care stay (<5 days/>5 days), functional warm ischaemic time (<20 mins/>20mins, <30mins), cold ischaemia time (<8 hours/>8 hours, <12 hours) and steatosis (<10%/>15%) [4]. This has resulted in up to 20% of donation after brain death (DBD) organs not meeting the clinical criteria [5] and a 78% increase in the discard rate of donation after circulatory death (DCD) livers [6]. These strict criteria can lead to a number of otherwise transplantable organs to being discarded [7]. Therefore, identifying specific pre-transplantation markers of liver damage could assist in expanding the pool of transplantable livers.

Currently, liver function tests are the standard assessments for establishing liver dysfunction, as evaluated by elevated concentrations of liver-enzymes such as alkaline phosphatase (ALP), alanine aminotransferase (ALT), aspartate aminotransferase (AST) and gamma-glutamyl transferase (GGT) [8, 9]. However, such tests lack sensitivity and specificity and can be affected by patient factors such as genetics, medicines and other non-associated diseases [10-13]. Thus far, transcriptomics and genomics have been used to discover biomarkers in live pathophysiology [14]. Metabolomics has also been employed to decipher metabolic fluxes in liver disease [15].

The objective of this study was to employ a molecular phenotyping approach to investigate hundreds of polar metabolites, at both pre- and post-transplantation, in hepatic tissue from two distinct donor types: *viz*. DBD and DCD. Following this, the association between metabolites that were different between donor types and clinical outcomes *viz*. early allograft dysfunction (EAD) and IGF were investigated. Then, prediction of IGF was calculated and survival analysis with metabolites and clinical variables. The study workflow is illustrated in **Fig. S1**.

## Materials and Methods

### Patients and samples

This study received prior approval from the ethics committee at King’s College Hospital, and informed consent was obtained from all subjects. The methods were carried out in accordance to the ethical guidelines of the 1975 Declaration of Helsinki. Overall 94 Tru-Cut tissue biopsies were obtained from liver allografts pre- and post-transplantation. The first (pre-transplant) biopsy was taken at the end of cold preservation, prior to implantation, and the second (post-transplant) biopsy was obtained approximately 1 hour after graft reperfusion. A separate biopsy was obtained for histopathological evaluation of donor steatosis. Biopsies were immediately snap-frozen in liquid nitrogen and stored at −80°C until extraction for LC-MS analysis. In all procedures, liver allografts were flash-cooled and perfused with University of Wisconsin preservation solution until the time of transplantation.

The study included two types of adult donors: DBD (n=35) and DCD (n=12). A wide spectrum of donor clinical data was collected for comparison among groups and for correlation with metabolite levels. In the DCD group, Functional WIT was calculated from the time when systolic blood pressure was below 50 mmHg to the time of cold perfusion. Total WIT is the sum of Functional WIT, Hepatectomy time (time from cold perfusion to hepatectomy) and Bench perfusion time (time from bench perfusion to liver graft into icebox). All recipients were patients with stable chronic liver disease who did not require hospitalization prior to transplantation. After transplantation, all patients received immunosuppressive therapy with tacrolimus and prednisolone. Graft performance was assessed based on AST, INR and bilirubin levels after transplantation [16]. According to graft performance, recipients were classified into two groups: showing EAD (n=10) and IGF (n=37). The survival data was collected for 34 recipients from the time of transplantation (between 2011 and 2013) till April 2019. The relevant donor and recipient details are listed in **Table 1**.

**Table 1.** Demographic characteristics and clinical data for 94 subjects involved in this study.

| Donor |  | DBD (N=35) | DCD (N=12) | p-value<sup>[b]</sup> |
| --- | --- | --- | --- | --- |
| Age(years) |  | 53(25-82) | 56(35-76) | 0.526 |
| Gender(female/male) |  | 19/16 | 6/6 | 1 |
| Hepatic steatosis | No | 14 | 7 | 0.305 |
|  | Mild (<30%) | 18 | 3 |  |
|  | Moderate (30-60%) | 3 | 2 |  |
| GGT (IU/L) <sup>[a]</sup> |  | 52(6-208) | 92(21-315) | 0.342 |
| AST (IU/L) <sup>[a]</sup> |  | 85(22-517) | 161(15-392) | 0.139 |
| ALT (IU/L) <sup>[a]</sup> |  | 72(12-268) | 97(13-201) | 0.623 |
| Bilirubin (μmol/L) <sup>[a]</sup> |  | 11(3-37) | 12(4-26) | 0.695 |
| ITU stay (days) |  | 4(1-28) | 4(1-10) | 0.168 |
| Inotrop support (Y/N) |  | 19/16 | 6/6 | 1 |
| Functional WIT (min) |  |  | 21(9-33) |  |
| Hepatectomy time (min) |  |  | 31(13-57) |  |
| Bench perfusion (min) |  | NA | 28(10-44) | NA |
| Total WIT (min) |  |  | 79(46-100) |  |
| CIT (min) |  | 504(210-840) | 457(270-720) | 0.212 |
| Recipient |  | DBD (N=35) | DCD (N=12) | p-value<sup>[b]</sup> |
| Age (years) |  | 44(20-65) | 54(46-70) | 0.029 |
| Gender (female/male) |  | 13/22 | 5/7 | 1 |
| BMI (kg/m<sup>2</sup>) |  | 25.8(18.4-34.6) | 27.3(22.1-35.8) | 0.277 |
| MELD Score (median) |  | 12.5 | 11.5 | NA |
| ALD |  | 9 | 3 | NA |
| PSC |  | 5 | 0 |  |
| HCV |  | 1 | 2 |  |
| HCC |  | 1 | 2 |  |
| BA |  | 0 | 0 |  |
| Others |  | 19 | 5 |  |
| AST (IU/L) <sup>[a]</sup> |  | 480(10-7485) | 613(18-5307) | 0.494 |
| Bilirubin day 7 (μmol/L) |  | 56(7-258) | 52(12-103) | 0.772 |
| INR day 7 |  | 1.04(0.85-1.21) | 1.06(0.92-1.3) | 0.909 |
| EAD/IGF |  | 6/29 | 4/8 | 0.251 |
| Censored/Dead <sup>[c]</sup> |  | 22/3 | 7/2 | NA |

DBD, donation after brain death; DCD, donation after circulatory death; GGT, gamma-glutamyl transferase; AST, aspartate aminotransferase; ITU, intensive therapy unit; WIT, warm ischaemia time; CIT, cold ischaemia time; BMI, body mass index; MELD, model for end-stage liver disease; ALD, alcoholic liver disease; PSC, primary sclerosing cholangitis; HCV, hepatitis C virus; HCC, hepatocellular carcinoma; BA, biliary atresia; EAD, early allograft dysfunction; IGF, immediate graft function. Continuous values are expressed as means (minimum-maximum); NA, not applicable. Total WIT is the sum of Functional WIT, Hepatectomy time and Bench perfusion. <sup>[a]</sup> Tested on the day of operation, <sup>[b]</sup> Mann Whitney test (2-sided) or Fisher exact test (2-sided), <sup>[c]</sup> survival information was collected for 34 recipients.

### Sample treatment

Sample preparation for all 94 biopsies followed our previously published method [17]. 100 µl of the lower aqueous phase from all samples were transferred to clean vials for further analysis. Samples were kept in the chamber with targeted temperature of 4 °C, and the injection volume is 5 µl with full loop function (20 µl loop size). Chromatographic and spectrometric conditions for the analysis of polar metabolites was conducted according to published protocol [18]. QCs were run in between every 8 samples in random orders.

### Data analysis

All data was processed within ‘XCMS’ package in ‘R Studio’ (version 1.0.153), and the multivariate analyses were conducted in both ‘R Studio’ and ‘SIMCA’ (version 14, MKS Umetrics AB, Sweden). Multivariate analysis included pre- and post-transplant matched samples n=94 (DBD n=70, DCD n=24) and 17 QCs. Principle component analysis (PCA) was carried out to detect outlier(s) and to examine the distribution of QCs. All pre-transplant data was then divided into training dataset (DBD n=30, DCD n=5) and test dataset (DBD n=5, DCD n=7). Orthogonal projections to latent structures-discriminant analysis (OPLS-DA) model was built based on the training dataset to examine the profiling of pre-transplant samples in DBD and DCD groups. Test dataset was utilised to assess the prediction ability of the built model. S-plot derived from the OPLS-DA model was then applied to select features based on covariance p[1] and correlation p(corr) value (p[1]>0.1, p(corr)>0.4 and p[1]<-0.1, p(corr)<-0.4).

Metabolic features were measured in the LC-MS data using Waters MassLynx software (Waters Corporation, Milford, MA). Feature concentrations were expressed as ratios of peak areas to internal standard peak area. The identification was performed by using metabolites mass to search against in-house and public metabolite databases [19-21]. Their structure and fragmentation patterns in the MS2 data were studied by comparing with pure standard molecules.

To compare between DBD and DCD as well as between EAD and IGF groups at pre- and post-transplantation stages, levels of the identified metabolites and their ratios to one metabolite of all the other selected metabolites were explored and examined with univariate non-parametric Mann-Whitney test (2-sided) and Benjamini-Hochberg test. Furthermore, random forest machine learning and receiver operating characteristic (ROC) were applied to choose the best predictors of IGF from the above selected ratios. Three ROC curves were reported: the highest possible AUC with combination of clinical variables, metabolites only (or their ratios) and the highest AUC combination of metabolites and clinical variables. (package “caret”, “randomForest”, “pROC”, and “ggplot2” in R studio 1.0.153). To follow this, correlation analyses between annotated metabolites and clinical features (AST, Bilirubin, GGT) were conducted. Calculations were conducted in SPSS 23 (IBM: Armonk, United States). Figures were plotted in GraphPad Prism 6 (GraphPad, US). Metabolite ratios, clinical variables and the type of liver donor information were compared for their predictive power of the survival. Two logistic regression models were fitted to make predictions based on metabolite ratios and clinical variables, respectively. Third, the group variable was used for the predictions as such. Participants were stratified into two equal-sized groups based on each of the three prediction models, and survival of these strata were compared with Kaplan Maier curves. (package “survival”, “survminer”, and “ggplot2” in R 3.4.2).

## Results

### Clinical outcomes

Patient demographics for all 94 samples in both groups are represented in **Table 1**. There were no significant differences between DBD and DCD groups in age, EAD/ IGF, liver enzyme levels, hepatic steatosis or serum bilirubin levels. Differences were observed in recipient ages (*p*<0.05) between groups.

### Multivariate model and feature selection

The unit variance (UV) scaled dataset was first inspected for detection of outlier(s). Next, the comparison between DBD and DCD samples at pre-transplant stage was performed. An OPLS-DA model was built with a training dataset (DBD n=30, DCD n=5), and the model was tested with a test dataset (DBD n=5, DCD n=7). As shown in the misclassification table, the test samples in DBD group can be predicted with 100% accuracy, while the DCD samples were predicted with 85.71% accuracy (**Table S1**).

In order to identify which metabolic features were the strongest discriminators between DBD and DCD at pre-transplant, an S-plot (**Fig. S2**) derived from the OPLS-DA model was used to select 12 features on the criteria stated in the method section. From the 12 selected features, 5 metabolites were annotated (**Table S2)**.

Five features were identified as purines at pre-transplant and their levels between DBD and DCD were represented as bar plots in **Fig. S3**. Additionally, jittered scatterplots representing the ratios of four purines to urate, illustrated in **Fig. 1**, were plotted. At pre-transplant stage, the ratio of AMP/urate, adenosine/urate, adenine/urate and hypoxanthine/urate were significantly higher in the DBD group compared to DCD (*q*<0.001). Moreover, the scatter plots showed that the ratio of AMP/Urate and adenine/urate were higher in the IGF group compared to EAD. The Mann-Whitney test confirmed that the mean ratios of adenine/urate and AMP/urate were significantly different between EAD and IGF (*q*<0.05). Adenosine/urate and hypoxanthine/urate showed no significant difference in the distribution between EAD and IGF groups.

**Fig. 1.**
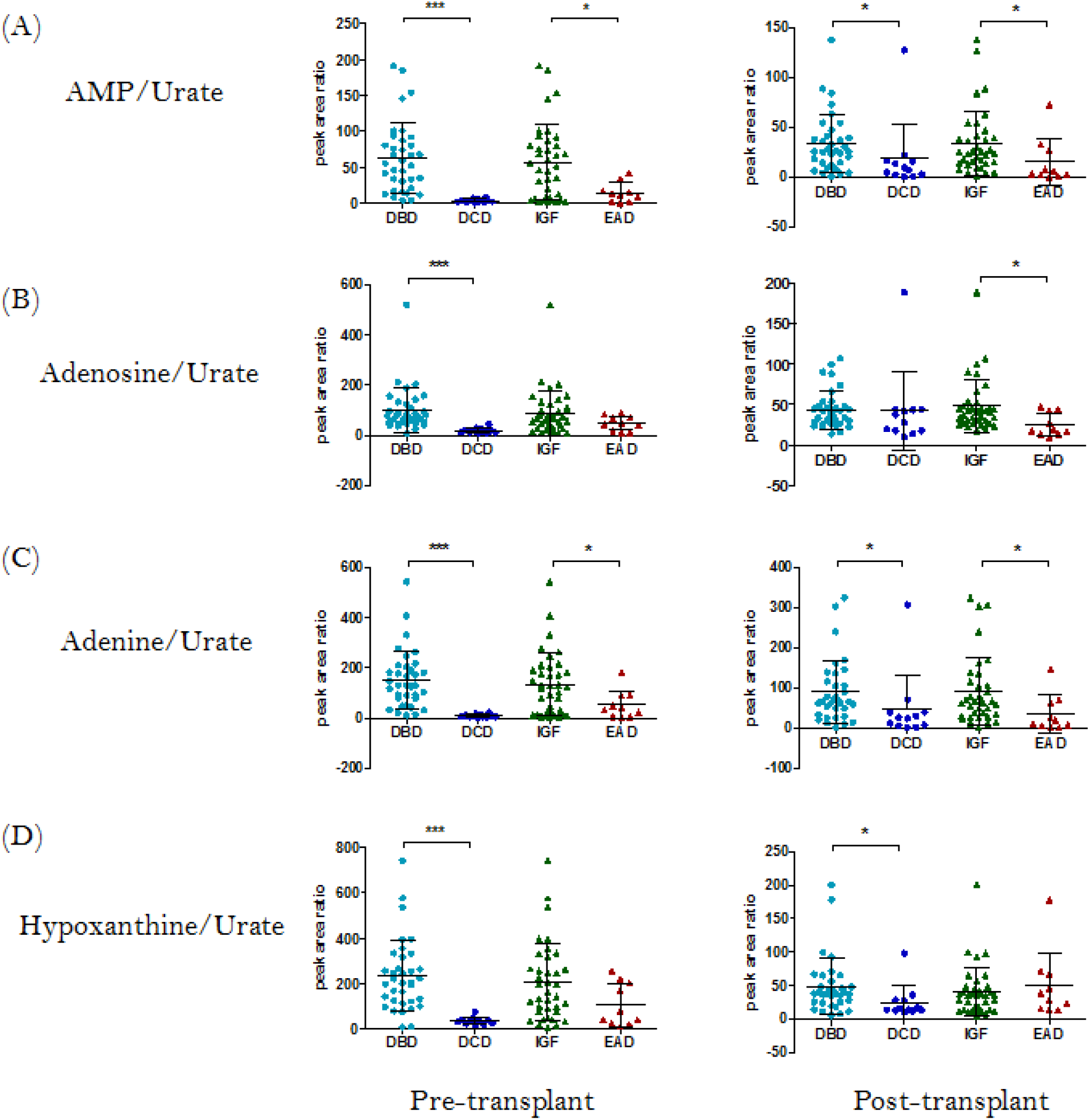
Jittered scatter plots of 4 ratios in four groups at both transplant stages. (A) AMP/Urate. (B) Adenosine/Urate. (C) Adenine/Urate. (D) Hypoxanthine/Urate. AMP, adenosine monophosphate; DBD, donation after brain death; DCD, donation after circulatory death; IGF, immediate graft function; EAD, early allograft disfunction. Results represented as mean ± SD, p-value was derived from Mann-Whitney test, followed by Benjamini-Hochberg FDR correction (\**q*<0.05, \*\**q*<0.01, \*\*\**p*<0.001).

At the post-transplant stage, AMP/urate, adenine/urate and hypoxanthine/urate (*q*<0.05) were significantly higher in the DBD group compared to DCD. Additionally, the scatter plot illustrated that the ratios of AMP/urate, adenosine/urate, and adenine/urate were elevated in the IGF group compared to EAD (*q*<0.05).

### Random forest with metabolites and clinical variables

Machine learning was applied to investigate variables to act as classifiers between the EAD and IGF groups. From the included variables (AMP/urate, adenine/urate, hypoxanthine/urate, adenosine/urate, ALT, bilirubin, AST, GGT, steatosis status and donor age), high importance scores for IGF were observed from the ratios of AMP/urate, adenine/urate, hypoxanthine/urate and ALT (**Fig. 2A)**. The prediction ability of purine ratios and ALT at pre-transplant were evaluated with ROC analysis. The accuracy, area under the curve (AUC), sensitivity and specificity for individual metabolites, enzymes, as well as various combinations in predicting IGF are listed in **Table 2**. The combination of the three purine ratios and ALT showed reliable prediction ability with high AUC, while the combination of four purine ratios demonstrated relatively higher accuracy, specificity and sensitivity value (**Fig. 2B**). Using random forest, a panel composed by ALT and AMP, adenine, hypoxanthine ratio to urate predicted IGF with AUC 0.84 (95% CI [0.71, 0.97]). In comparison AUC 0.71 (95%CI [0.52, 0.90]) was achieved by the clinical measures.

**Table 2.**
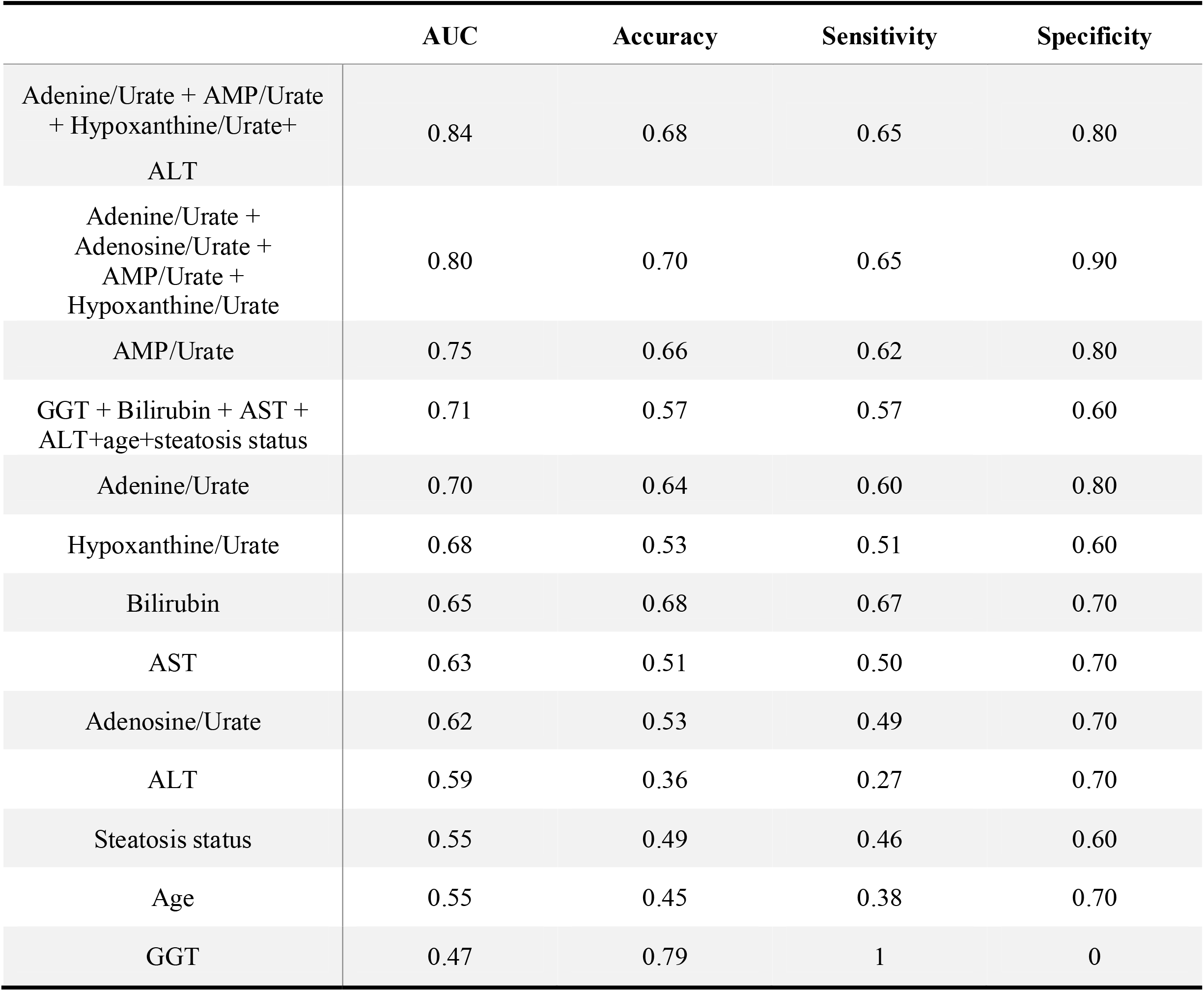
ROC analysis for 5 annotated metabolites and 5 donor clinical parameters at pre-transplant for the prediction of IGF.

**Fig. 2.**
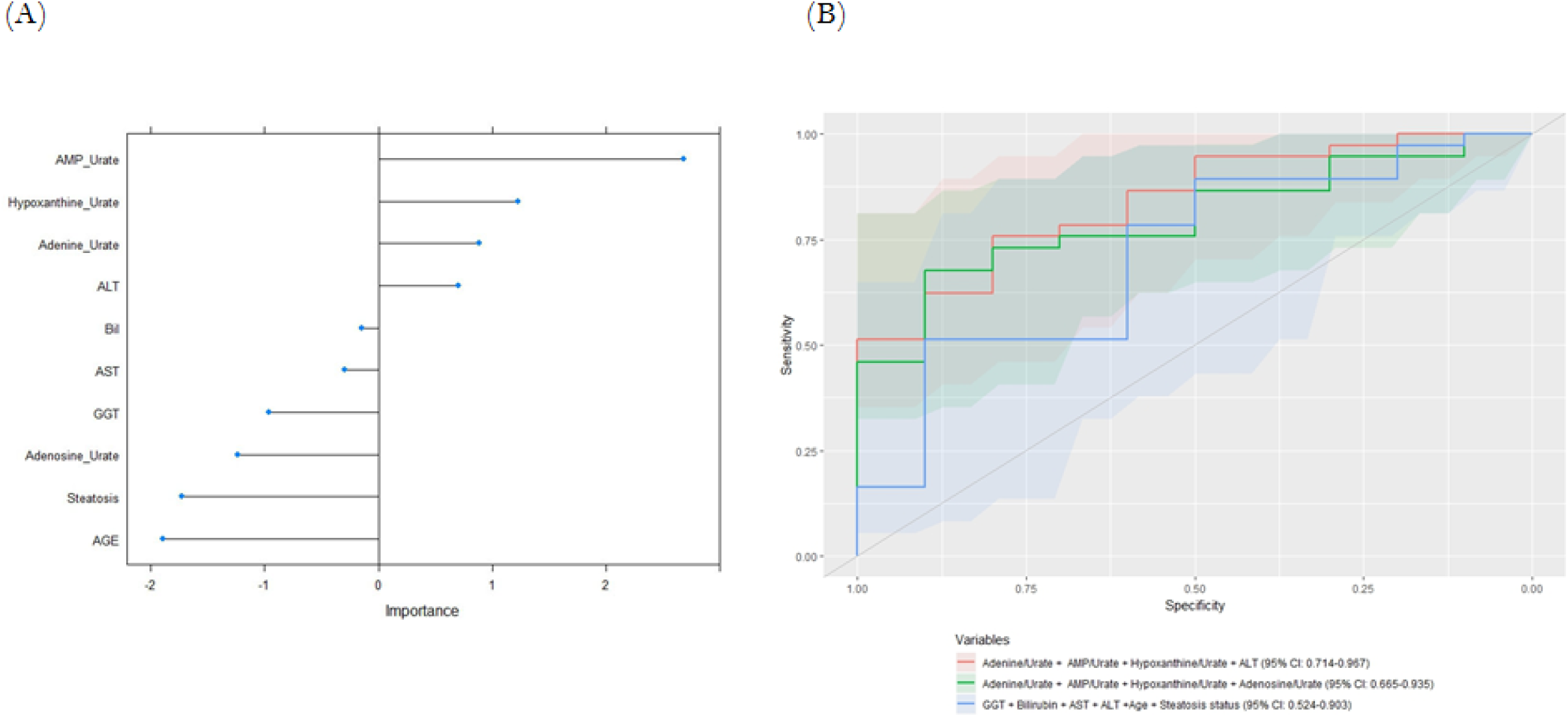
(A) Variable importance plot derived from the random forest model. (B) ROC curve prediction of IGF based on highest AUC with the combination of clinical variables, metabolite ratios only and combination of metabolites and clinical variables.

In order to investigate whether liver enzymes were associated to these purines, partial correlation analysis was performed. Purine relative amounts in pre- and post-transplant samples, together with AST, Bilirubin and GGT in donors on the day of operation (day 0) and in recipients on the day after operation (day 1) were included for correlation analyses. In **Table 3**, the only significant correlation (*q*<0.05) was observed between hypoxanthine and bilirubin after Benjamini-Hochberg correction.

**Table 3.**
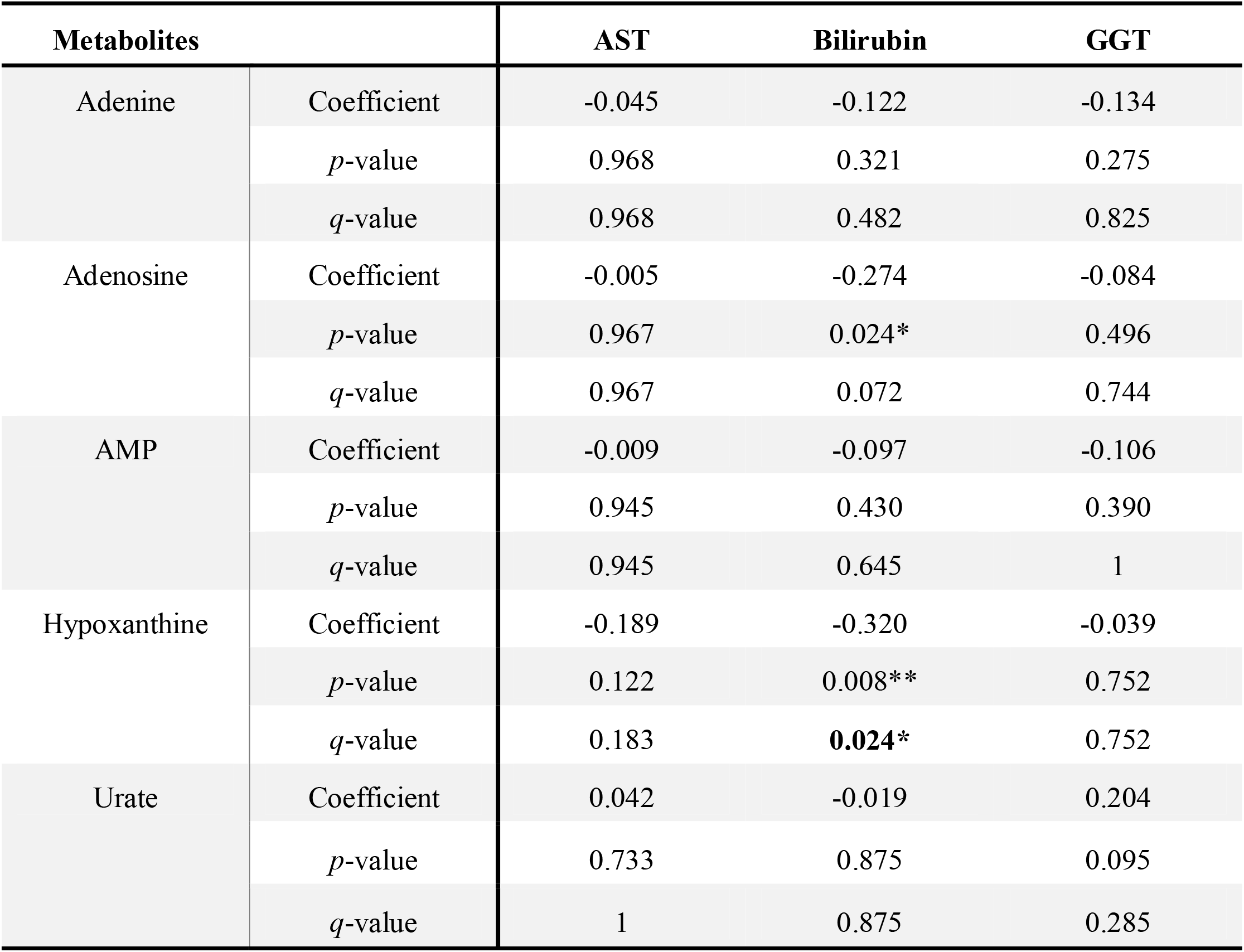
Partial correlation analysis (Pearson’s correlation with adjusting for patient age) between the 5 selected metabolites and liver enzymes. *p*-values were represented as *q*-values after applying Benjamini-Hochberg correction. \**p* or *q*<0.05, \*\**p* or *q*<0.01.

| Metabolites |  | AST | Bilirubin | GGT |
| --- | --- | --- | --- | --- |
| Adenine | Coefficient | -0.045 | -0.122 | -0.134 |
|  | *p*-value | 0.968 | 0.321 | 0.275 |
|  | *q*-value | 0.968 | 0.482 | 0.825 |
| Adenosine | Coefficient | -0.005 | -0.274 | -0.084 |
|  | *p*-value | 0.967 | 0.024* | 0.496 |
|  | *q*-value | 0.967 | 0.072 | 0.744 |
| AMP | Coefficient | -0.009 | -0.097 | -0.106 |
|  | *p*-value | 0.945 | 0.430 | 0.390 |
|  | *q*-value | 0.945 | 0.645 | 1 |
| Hypoxanthine | Coefficient | -0.189 | -0.320 | -0.039 |
|  | *p*-value | 0.122 | 0.008** | 0.752 |
|  | *q*-value | 0.183 | **0.024\*** | 0.752 |
| Urate | Coefficient | 0.042 | -0.019 | 0.204 |
|  | *p*-value | 0.733 | 0.875 | 0.095 |
|  | *q*-value | 1 | 0.875 | 0.285 |

### Survival analysis with purines, clinical variables and donation groups

The purine ratio predictor (AMP/urate, adenine/urate, hypoxanthine/urate, adenosine/urate) stratified the participants completely: all five deaths occurred in the <50% strata, in which the metabolites predicted a lower chance of survival (**Fig. 3A**; p = 0.073). For the clinical predictor (ALT, bilirubin, AST, GGT, steatosis status and donor age), three out of the five deaths occurred in the <50% strata, which indicates no significant prediction (**Fig. 3B**; p = 0.54). Similarly, three out of five deaths occurred in the DBD group, group class could not predict survival (**Fig. 3C**; p = 0.15).

**Fig. 3.**
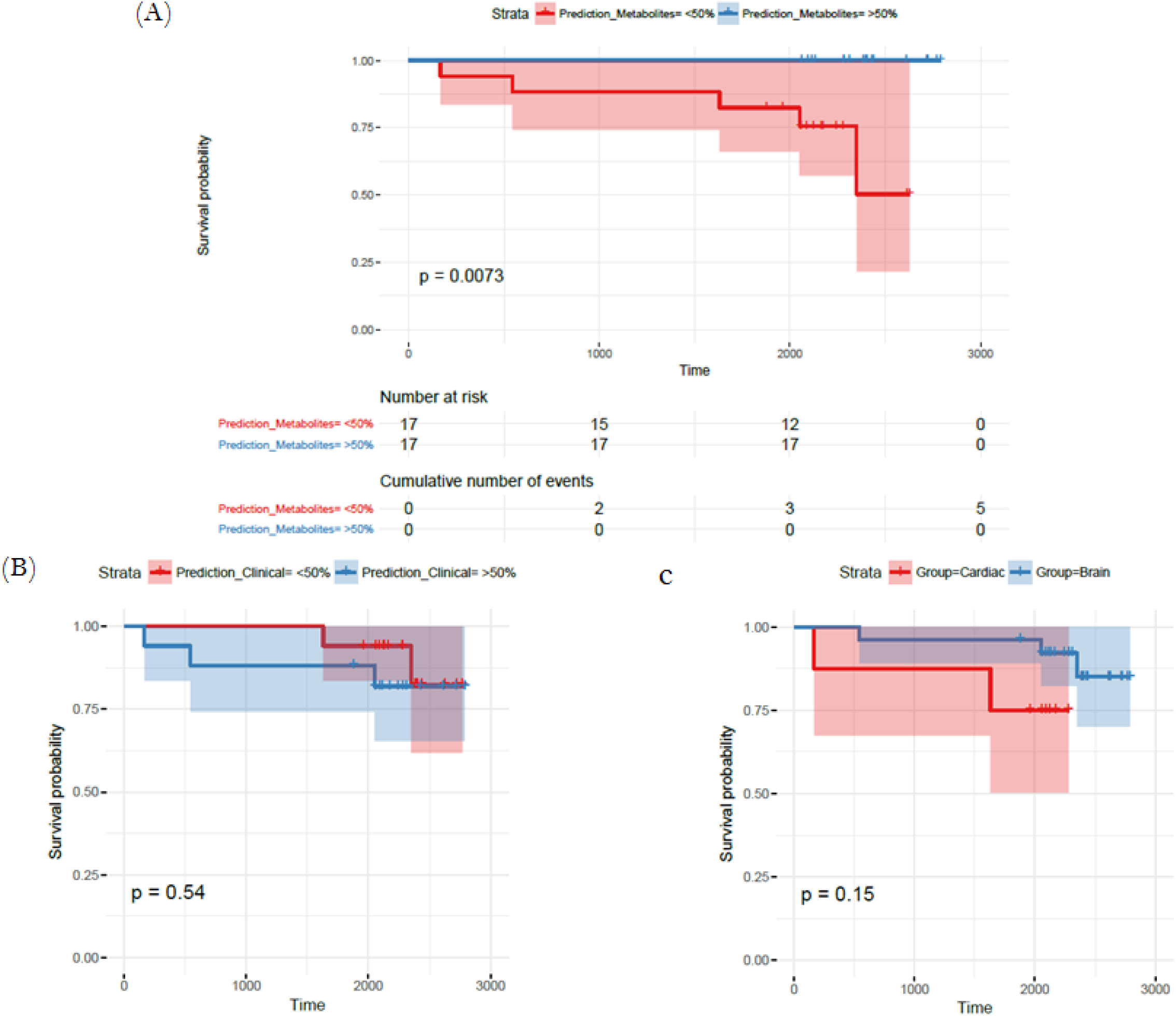
Kaplan-Meier plots of time estimates survival of patients with three different groups od predictors. (A) Metabolite ratios as predictors. (B) Clinical variables as predictors, c) donation groups as predictors.

## Discussion

The five metabolites that were highly correlated to DCD are generated from the purine metabolism pathway (**Fig. 4**) [22, 23]. Metabolites in the purine pathway have a myriad of functions and are important in regulating inflammation [24], in oxidative injury and as markers of cell death. In liver tissue undergoing cold and warm ischemia, the dysregulation of their levels could be related to energy, inflammation and ischemic tissue damage [25-27].

**Fig. 4.**
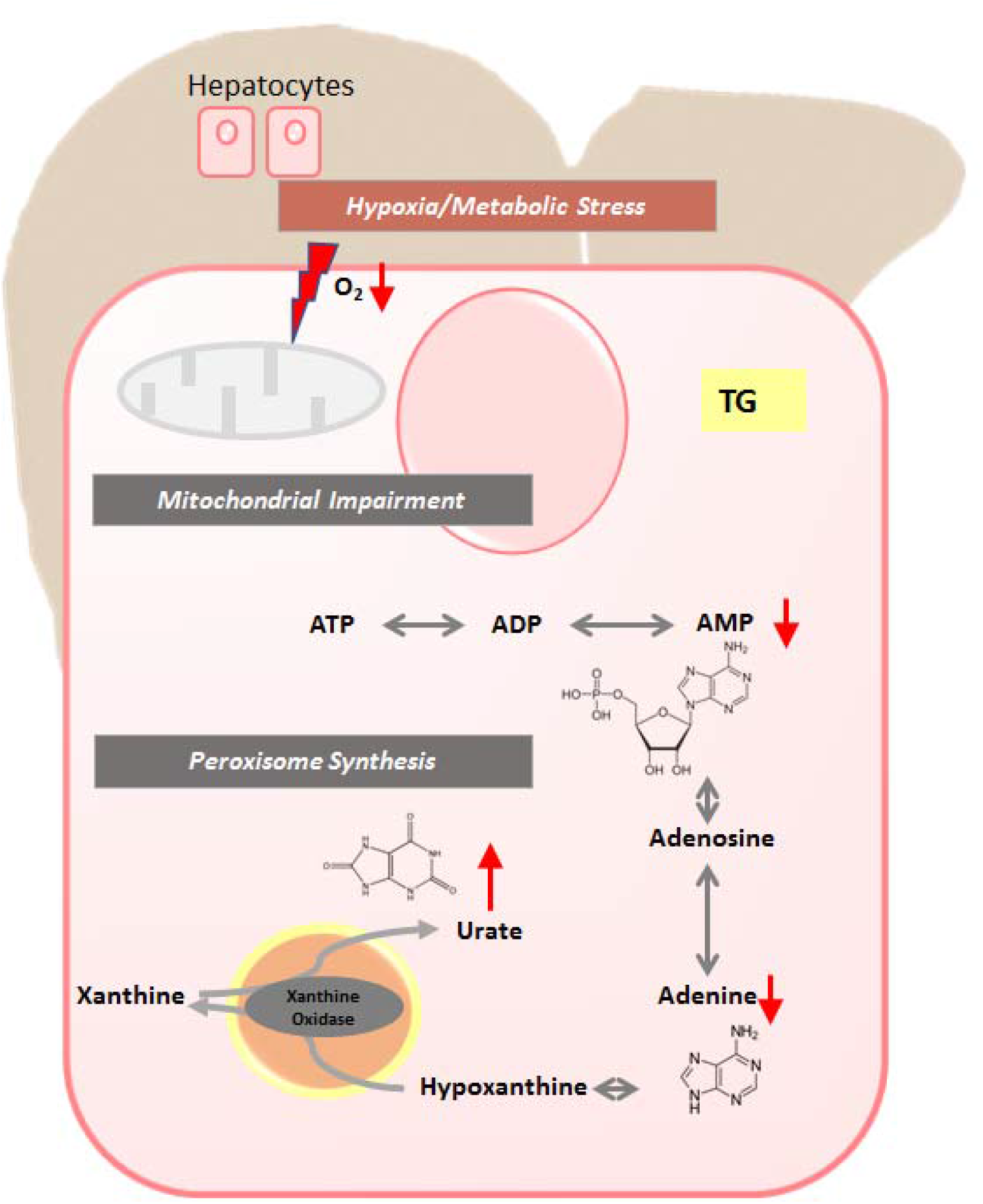
The proposed metabolic changes taking place in an explanted liver. During energy production, phosphate groups are sequentially hydrolysed from ATP, creating ADP then AMP. From AMP the other metabolites are generated via a number of catabolic pathways. ATP, adenine triphosphate; ADP, adenine diphosphate; AMP, adenine monophosphate.

Purines can act as physiological regulators of leucocyte function [28], but to be functional they must be released in appropriate loci following stimuli [29]. It is thought that liver inflammation is due to a cascade of inflammatory events that occur mainly in the donors after brain death [30]. On the other hand, our studies have shown that DCD grafts undergo low inflammation and increased hepatocellular damage due to warm ischaemia time [30, 31].

In this study AMP and adenine were found critically decreased in DCD. Studies have shown that adenine and AMP have a protective function during ischaemia [32]. In addition to being mediators for graft recovery, Roy *et al*. found that having high levels of AMP during ischaemia when oxygen is low, indicates ATP is still being generated [33, 34]. This might explain why DBD allografts and the IGF group showed increased levels of both metabolites, as a higher energy reserve could improve the post-transplant graft function [35].

AMP is also known to be protective during inflammation. It is converted from ATP and ADP by ectoapyrase (CD39) and released at the site of vascular injury when platelets aggregate to promote endothelial barrier function during inflammation [23]. Michael *et al*. found that an overexpression of CD39 and hence AMP conferred protection in both warm and cold hepatic ischaemia [36].

Adenine has been employed as a substrate during recovery. It has been shown that dying cells as a result of ischaemia undergo lysis to release adenine [37]. Kartha *et al*. demonstrated *in vitro* that adenine nucleotides accelerated structural and functional recovery in epithelial cells [38]. This would suggest that DBD liver allografts (**Fig. S3B**) with elevated levels of adenine at pre-transplantation might be able to recover sooner.

Although not significantly elevated at pre-transplantation, adenosine and hypoxanthine, showed the same trend as AMP and adenine. Wyatt *et al*. found that a solution containing hypoxanthine and adenosine enhanced functional organ recovery after I/R injury in dogs [39].

Increased levels of urate were observed in DCD livers at pre-transplant (*q*<□0.001) (**Fig. S3E**). In humans, urate is the final product of purine metabolism [40]. In an experiment conducted by Matthew *et al*., in which hepatic ischaemia was induced for 30 min followed by 60 min of reperfusion. After ischaemia, urate levels had increased by over 300% and during the first 30 min of reperfusion by 600% [41]. Clear differences were revealed between DBD and DCD, as well between IGF and EAD groups when the ratios to urate were investigated (**Fig. 1**). Epidemiological studies have also suggested that during IRI urate levels are increased [42]. DCD allografts are more prone to IRI due to being exposed to the period of warm ischaemia [43].

We wanted to calculate the prediction of classifiers including purines for outcomes of IGF and longer-term survival. The model for IGF revealed that the diagnostic potential of combination of three ratios (AMP/urate, adenine/urate and hypoxanthine/urate) and ALT was the top IGF prediction at 84% with a confidence interval from 71% to 96%. While higher accuracies were observed when purines were compared to known risks and enzyme makers (**Table 2**), the confidence interval shows that our study needs a replication in a bigger cohort. The survival analysis revealed that metabolite ratios were the best predictor of survival, compared to classifiers with clinical variables and the type of the liver donor. Again, metabolite ratios predicted the deaths in the small data set in comparison to clinical variables and the type of donor. These preliminary results indicate that purine ratios may predict prognosis beyond the clinical profile or the type of liver donation. However, we reiterate that the small number of events in the current study and a population that was limited to biopsies from operations conducted in one centre warrants that validation will be planned as a multicentre trial assessing graft function.

In this study, the combination of AMP/urate, adenine/urate, hypoxanthine/urate and ALT proved to have higher prediction ability compared to a combination of conventional liver function and risk markers. This study proposes a panel of small molecules at pre-transplantation that can aid testing liver tissue quality for liver transplantation.

## Data Availability

The datasets generated during and/or analysed during the current study are available from the corresponding author on reasonable request.

## Supporting information

Additional Supporting Information may be found online in the supporting information tab for this article.

**Supplementary Table S1**. Misclassification table for the test dataset based on the training dataset model

**Supplementary Table S2**. Annotation of markers based on molecular weight, retention time and collision induced dissociation fragmentation of 5 metabolites.

**Supplementary Fig. S1**. Study workflow. PCA, principle component analysis; OPLS-DA, orthogonal projections to latent structures-discriminant analysis; EAD; early allograft dysfunction; IGF, immediate graft function; ROC, receiver operating characteristic.

**Supplementary Fig. S2**. Metabolic feature selection from the S-plot. All the dots represent detected features, and the pink dots were selected for annotation.

**Supplementary Fig. S3**. Bar plots of 5 metabolites in four groups at both transplant stages. (A) AMP, (B) Adenosine, (C) Adenine, (D) Hypoxanthine and (E) Urate.

